# Within-host genetic diversity of SARS-CoV-2 in the context of large-scale hospital-associated genomic surveillance

**DOI:** 10.1101/2022.08.17.22278898

**Authors:** Alexandra A. Mushegian, Scott W. Long, Randall J. Olsen, Paul A. Christensen, Sishir Subedi, Matthew Chung, James Davis, James Musser, Elodie Ghedin

**Affiliations:** Systems Genomics Section, Laboratory of Parasitic Diseases, National Institute of Allergy and Infectious Diseases, National Institutes of Health, Bethesda, MD USA; Laboratory of Molecular and Translational Human Infectious Diseases Research, Center for Infectious Diseases, Department of Pathology and Genomic Medicine, Houston Methodist Research Institute and Houston Methodist Hospital Houston, Texas, 77030; Division of Data Science and Learning, Argonne National Laboratory, 9700 S. Cass Ave., Lemont, Illinois, 60439; University of Chicago Consortium for Advanced Science and Engineering, 5801 South Ellis Avenue, Chicago, Illinois, 60637

## Abstract

The COVID-19 pandemic has resulted in extensive surveillance of the genomic diversity of SARS-CoV-2. Sequencing data generated as part of these efforts can also capture the diversity of the SARS-CoV-2 virus populations replicating within infected individuals. To assess this within-host diversity of SARS-CoV-2 we quantified low frequency (minor) variants from deep sequence data of thousands of clinical samples collected by a large urban hospital system over the course of a year. Using a robust analytical pipeline to control for technical artefacts, we observe that at comparable viral loads, specimens from patients hospitalized due to COVID-19 had a greater number of minor variants than samples from outpatients. Since individuals with highly diverse viral populations could be disproportionate drivers of new viral lineages in the patient population, these results suggest that transmission control should pay special attention to patients with severe or protracted disease to prevent the spread of novel variants.

## Introduction

During the COVID-19 pandemic, emerging variants of SARS-CoV-2 have been globally tracked due to the rapid acquisition and sharing of whole genome sequence data^1^. As of July 2022, close to 12 million SARS-CoV-2 consensus genome sequences have been deposited to the GISAID repository (https://www.gisaid.org). These sequences represent summaries of petabytes of raw sequencing data, which cover the 30Kb RNA genome of SARS-CoV-2 at high redundancy. These deep sequence data could potentially be a rich source of information about the emergence of mutations in the virus population within the infected host, prior to transmission. Because an infected host is a dynamic, heterogeneous environment in which viruses replicate and compete under immunological pressure, it is of great interest to understand how much heterogeneity in the within-host viral population lies beneath the consensus viral genome sequence.

Studies of within-host viral diversity—variously referred to in terms of minor variants, quasispecies, low-frequency variants, or intrahost single nucleotide variants (iSNVs)— infer the viral diversity within a specimen from the relative abundances of sequencing reads supporting polymorphic sites^2^. Such studies aim to capture de novo mutations acquired by the virus over the course of within-host replication, as well as mixed infections acquired through transmission of multiple lineages. In principle, this information could be used to help predict the emergence of novel variants, to identify sites under evolutionary selection, or to help track transmission^3^. It is also of interest to determine if patient characteristics, behavior, or differences in clinical care strategies influence the magnitude of viral diversity generated and maintained in individual patients or during transmission. For example, there is evidence that SARS-CoV-2 variants with multiple novel mutations have emerged in patients with protracted infections ^4^.

However, the existing studies also acknowledge that such inferences must be made cautiously. Within-sample read diversity can also be due to sample contamination, especially with aerosolized PCR product; biases generated during reverse transcription, PCR, enrichment, and library preparation steps; sequencing errors; and artefacts generated during bioinformatic processing and read mapping^5^. Well-established methods exist for accounting for these processes when it comes to assembling consensus sequences, but it is considerably more difficult to accurately quantify within-sample variation without making special efforts to counteract these sources of error. Therefore, it is crucial to develop best practices for inferring minor variant diversity from viral deep sequencing data, especially from opportunistic datasets generated with consensus genome sequences as the primary goal and not minor variant analysis.

We explored the feasibility of extracting actionable signals about within-host viral genetic diversity from the deep sequence data underlying consensus genomes by focusing on a cohort from the Houston Methodist Hospital System. A network of eight hospitals and an associated research institute serving a demographically diverse city of 7 million people, HMH began using high-output Illumina instruments to sequence all SARS-CoV-2 specimens coming through the system in December 2020. End-to-end processing of samples, from collection through read generation, occurred within the same set of facilities and protocols with a high level of technical standardization and automation. The resulting consensus sequences were deposited in GISAID and used to track epidemiological trends in Houston^6–8^. The dataset is unique in that it densely samples a large population over an extended period of time with rich patient metadata linked to samples. However, these same advantages come with challenges from the perspective of minor variant tracking: samples are processed approximately sequentially, not in controlled or randomized batches, and there is limited opportunity in an active high-throughput sequencing facility of this scale to sequence technical replicates.

Previous studies of minor variants that carefully addressed sources of error and uncertainty have emphasized different aspects of within-host viral diversification^9–12^. Despite methodological differences, several broad observations have been consistent across studies: within-host diversity is generally low, albeit with some outlier samples containing high diversity; and within-host mutations apparently independently recur frequently between samples, impeding attempts to use minor variant information to infer transmission. We used our large dataset to delve more deeply into unanswered questions surrounding these observations. First, we used the patient metadata available to explore whether there are any patient characteristics associated with high minor variant richness, since such individuals might be disproportionate drivers of the emergence of new consensus mutations in an analogous way to a small number of patients driving superspreading events. Second, we examined a set of highly recurrent minor variants to investigate how many were systematic technical artefacts vs. hypermutable sites with potential phenotypic consequences. Theory and empirical data show that biases in de novo generation of mutations can skew evolutionary trajectories, with convergent traits often arising via pathways involving hypermutable sites.^13,14^ We find that while both phenotypically important mutations and probable artefacts regularly recur as minor variants, a robust association between high within-host virus diversity and patient hospitalization (admission into inpatient or ICU care) could be detected. The mechanism and direction of this association is unknown, but this observation supports the conclusion that transmission control in healthcare settings or from severely ill patients should be of particular focus in preventing the emergence of new variants.

## Results

### Sample inclusion criteria, minor variant detection and reproducibility

Between the beginning of December 2020 and the end of November 2021, a total 39,880 samples were collected at Houston Methodist, encompassing a wide range of symptomatic and asymptomatic patients and healthcare workers. These were sequenced across 70 NovaSeq sequencing runs. We narrowed down the dataset considerably according to various criteria such as the sequencing depth and the input quantity of viral RNA, approximated by the quantitative PCR cycle threshold value, Ct (see *Methods*). Since no-template negative controls were not sequenced, we used Ct>=40 samples as pseudo-negative controls to assess the level of background PCR amplicon contamination in each run, reasoning that background contamination of deeply sequenced samples does not necessarily impact consensus sequence calling but can affect the appearance of within-host diversity. We excluded runs containing at least three Ct>=40 samples in which the coverage breadth and depth were not statistically different from the Ct<40 samples in the run (t-test>=0.01). This conservative criterion resulted in the exclusion of 22 sequencing runs. We also limited all our analyses to samples with at least 100x coverage over at least 98% of the genome, and excluded samples where the consensus sequence was flagged as poor quality by Nextclade^15^ or that did not have a lineage assigned by Pango^16^, and used the earliest available sample from patients from whom multiple samples were collected. These initial filtering criteria narrowed the dataset to 15,389 samples with a lower average Ct value (19.96) than the total population (23.47) (**Fig.1a**).

**Figure 1.**
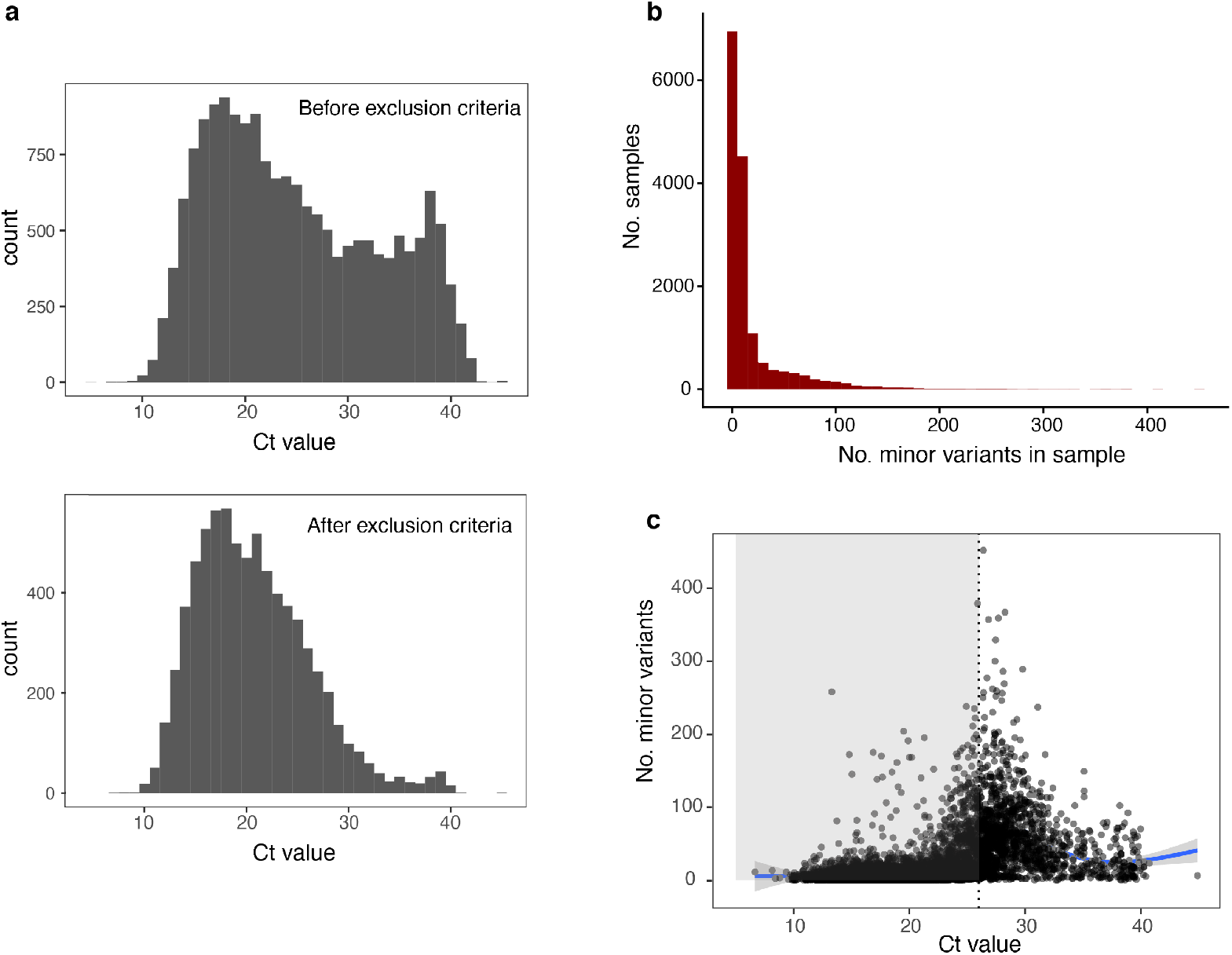
Distribution of minor variant richness in Houston Methodist SARS-CoV-2 specimens. **a**. Change in distribution of sample Ct values after applying minimum sequencing depth and background contamination exclusion criteria (see Methods). **b**. Number of minor variants per sample in 15,389 samples passing initial quality control criteria (minimum depth/breadth of coverage, run-level quality screening, consensus sequence assembly quality). **c**. Relationship between minor variant richness and sample input quantity (qPCR Ct value) in samples for which Ct values were available (n=7356). Vertical dotted line represents Ct<26, the threshold below which minor variants are generally reproducible.

Minor variants were identified using a variant calling pipeline (*timo*), which was previously demonstrated to have high precision and recall of minor variants, given a background rate of sequencing error^5^. The output of this pipeline also differs from many minor variant callers in that it identifies minor variants that are reversions to the ancestral reference allele at sites where the consensus allele differs from the reference. We considered minor variants at sites with a total depth of coverage of at least 100 reads, where the minority variant made up at least 1% of reads at the site at a minimum depth of 50 reads (thus requiring a more stringent standard of minimum minor variant frequency at sites with lower coverage). We excluded any minor variants at PCR primer binding sites of either of the primer sets used over the course of the study, and excluded any sites called as gaps or Ns in the consensus or minority fraction. For 54 of the samples, technical replicates re-sequenced from the original RNA were available, which we used to assess how many of the minor variants were reproducible. We found that both the presence/absence and within-host frequency of minor variants were highly reproducible in samples with Ct values <26 (**SuppFig.1 a,b**). This was consistent with the range of input quantities at which minor variants were reproducible in several previous studies^11,17^. At higher Ct values, many more minor variants failed to be detected in the second replicate of sequencing or were detected but at substantially different frequencies. Sequencing depth either across the whole genome or at individual sites was not clearly associated with reproducibility (**SuppFig.1 c,d**). We thus concluded that minor variants were more likely to be spurious in lower-input samples but were reliably detected in a single replicate of sequencing in higher-input samples, and that sample input amount rather than sequencing depth was a more reliable indicator of sample quality for this purpose.

### Within-host minor variant diversity

In the 15,389 samples passing the initial quality control criteria, 9,771 (63%) contained minor variants at fewer than 10 nucleotide positions, with a long tail of samples containing much higher diversity (**Fig.1b**). Consistently with previous studies^9^, there was a strong correlation with Ct value, with the most diversity concentrated in moderately low-input samples (Ct∼28-30) (**Fig.1c**). There could be technical or biological reasons for higher minor variant richness in lower viral load samples: they are inherently more variable due to stochastic sampling and thus are more sensitive to contamination and technical artefacts, but they also could have been collected early or late in the infection, reflecting different points in the mutation/selection trajectory of the viral population. Given the previous findings about reproducibility, we limited further analysis to samples with Ct<26, acknowledging that although minor variant data from these samples is likely more accurate, this stringent criterion likely affects the composition of the patient cohort studied, since it excludes patients with low viral loads. Because diagnostic PCR was carried out on different instruments across the healthcare system, Ct values were not available for all samples; thus the final dataset contained 6,140 samples. The median number of minor variants in samples in this dataset was 5, with a maximum of 379. A slight positive association with Ct value remained, which is likely due to genuine biological factors in this range of input values (**Fig.1b**, *grey region*).

The final dataset of samples spanned 47 sequencing runs encompassing several distinct stages of the epidemic. In December 2020 and early January 2021, the dominant consensus sequences were from variant B.1.2 and an assortment of smaller lineages. Starting in late January, a period of declining cases was strongly dominated by the Alpha variant (B.1.1.7), which was replaced in July by the Delta variant (B.1.617.2) in a late summer/autumn surge (**SuppFig.2**). Sample collection dates were roughly, but not completely, chronologically associated with runs (**SuppTable 1)**. Even after stringent filtering criteria, there remained a run-level effect on the detection of minor variants, which appeared to be related to the run-level average of sequencing depths rather than individual sample sequencing depths (**SuppFig.3**). Sequencing batch effects are therefore important to consider when assessing minor variant diversity.

### Clinical correlates of within-host diversity

Having controlled as much as possible for artefacts and systematic errors, we next determined if unusually high minor variant richness was associated with any clinical features. From available medical records, we obtained deidentified data on patient demographics (age group, sex, and ethnicity), comorbidities (chronic heart, lung, liver and kidney disease; hypertension; diabetes; obesity; HIV status; previous organ transplant status; cancer), and aspects of clinical treatment (whether the patient was hospitalized and/or was treated with plasma or monoclonal antibodies). Because many of these factors are highly correlated with each other, we used a random forest classification model to query the relative importance of these factors in grouping samples as “high diversity” or “low diversity.” We considered “high diversity” samples to be those with more than 5 minor variants, which was the median number. While the overall performance of the random forest model was poor (ROC AUC=0.58), suggesting that the clinical features included were insufficient to classify high vs. low diversity samples without additional information, the most important variable in the trained model was hospital admission. A chi-squared test confirmed that high-variant samples were overrepresented among hospitalized patients—treated as inpatients or admitted to intensive care—compared to outpatients (X^2^ = 131.58, df = 1, p< 2.2e-16). The odds ratio of the association of hospitalization with high minor variant diversity was 1.84 (95% CI 1.66-2.05). There was a higher proportion of hospitalized patients, as compared to non-hospitalized individuals, with at least 1 minor variant, more than 5, or more than 10 minor variants in the sample (**SuppFig.4**).

To take sample viral input amount and sequencing run batch into account, we constructed a linear mixed effects model with hospital admission and sample Ct value as fixed effects, sequencing run as a random effect, and the log-transformed number of minor variants in the sample as the response variable. Hospital admission and Ct value were both significantly associated with minor variant diversity (**Table 1a, Fig.2a**). One plausible reason samples from patients requiring hospitalization could have higher minor variant richness is if they were on average collected later in the infection than samples from non-hospitalized patients. We did not have information on the number of days post infection for any of the samples. As another way to probe the relationship between disease severity and minor variant richness, that is less likely to be related to infection duration, we examined minor variant diversity in samples from July 2021 onwards when an appreciable number of vaccine breakthrough cases occurred. Assuming that vaccination was associated with less severe disease even among the population not requiring hospitalization, we compared minor variant richness in the infections of fully vaccinated and unvaccinated or partially vaccinated non-hospitalized individuals. At comparable Ct values, minor variant diversity was significantly higher (p <0.05) in the unvaccinated than in the fully vaccinated cases (**Table 1b, Fig.2b**). Finally, we compared minor variant diversity in samples from healthcare workers undergoing voluntary surveillance testing with samples from non-hospitalized patients. In this case, minor variant richness was not significantly different between these two groups (**Table 1c, Fig.2c**).

**Figure 2.**
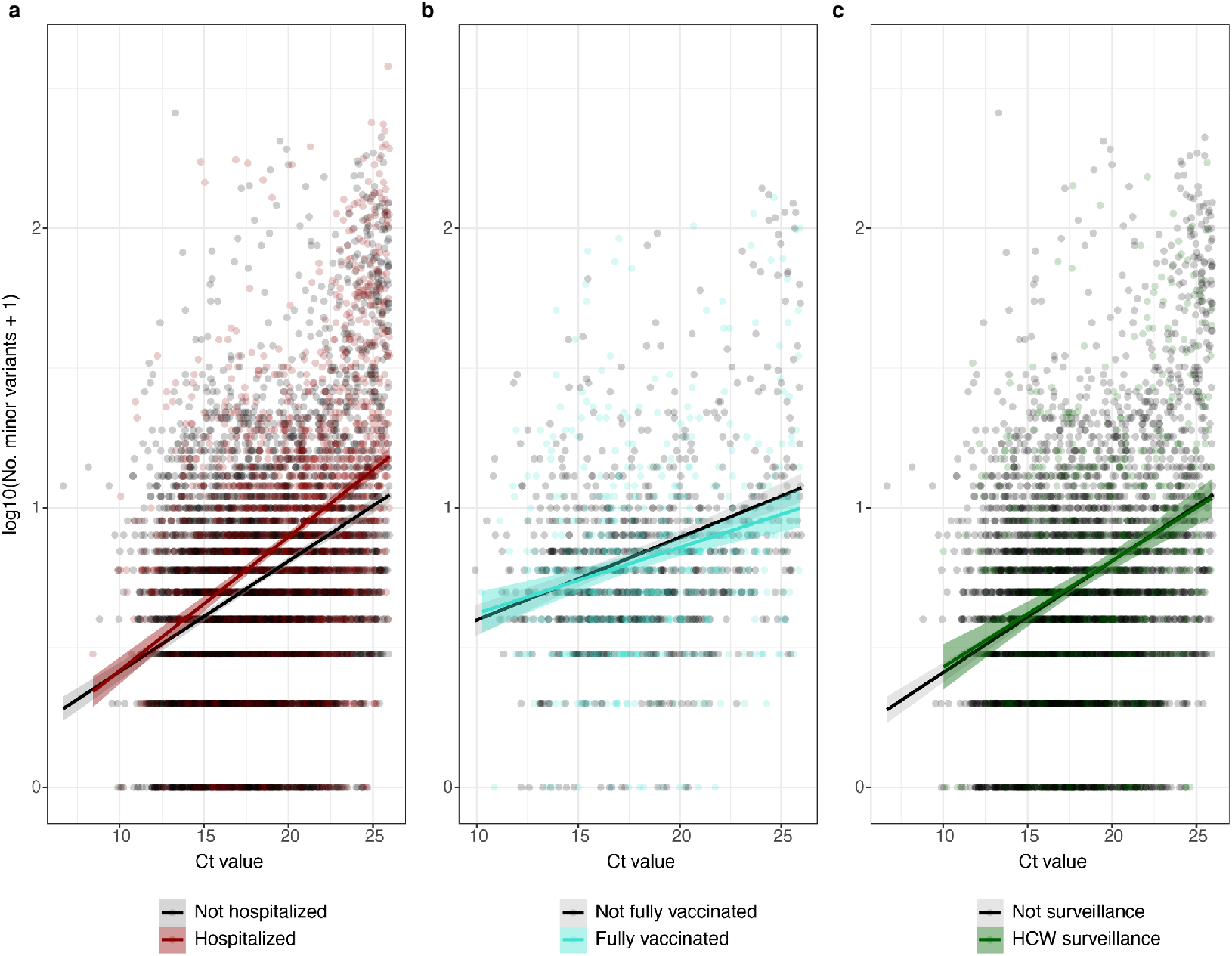
Association between patient status and SARS-CoV-2 diversity. **a**. Minor variant richness across Ct values in patients requiring hospitalization (inpatient or ICU) vs. outpatients (n=6140). **b**. Minor variant richness in fully vaccinated vs. unvaccinated or partially vaccinated patients, limiting samples to those collected in July 2021 or later and only from patients not requiring hospitalization (n=1366). **c**. Minor variant richness in non-hospitalized patients vs. healthcare workers undergoing voluntary surveillance (n=3874).

**TABLE 1.** Linear mixed-effects models for association of hospitalization, vaccination and healthcare worker surveillance samples on minor variant richness.

|  | numDF | denDF | F-value | p-value |
| --- | --- | --- | --- | --- |
| (Intercept) | 1 | 6090 | 602.4491 | <.0001 |
| Ct | 1 | 6090 | 1370.695 | <.0001 |
| admitted_hospital | 1 | 6090 | 77.6004 | <.0001 |
| Ct:admitted_hospital | 1 | 6090 | 11.5969 | 7.00E-04 |

|  | numDF | denDF | F-value | p-value |
| --- | --- | --- | --- | --- |
| (Intercept) | 1 | 1346 | 297.22177 | <.0001 |
| Ct | 1 | 1346 | 152.42258 | <.0001 |
| vaccine_status | 1 | 1346 | 4.52737 | 0.0335 |
| Ct:vaccine_status | 1 | 1346 | 4.31531 | 0.0380 |

|  | numDF | denDF | F-value | p-value |
| --- | --- | --- | --- | --- |
| (Intercept) | 1 | 3824 | 455.9225 | <.0001 |
| Ct | 1 | 3824 | 669.4242 | <.0001 |
| surveillance_sample | 1 | 3824 | 0.1950 | 0.6588 |
| Ct:surveillance_sample | 1 | 3824 | 0.0207 | 0.8857 |

As a complementary approach to evaluating the association between patient factors, sample characteristics, and minor variant richness, we constructed a LASSO regression model containing the comorbidities, treatments, and demographic factors, as well as Ct values, median sequencing depth, collection month and run. In the best model (lambda=0.0013) the deviance ratio was 0.226, meaning that the combination of variables we included explained approximately 22.6% of the variation in the log-transformed number of variants. We also constructed a version of this model which excluded all factors that had a strong temporal bias (vaccination status, consensus variant, and collection month), because temporal trends in this study design were impossible to separate from sequencing batch (run) effects. In this model (deviance ratio 0.17), hospital admission was clearly associated with the highest increase in minor variant richness (**SuppFig.5**).

Taken together, these complementary modeling strategies suggest there is substantial unexplained variation in within-host minor variant richness. They also highlight that severity of disease—as exemplified here by hospitalization or lack of vaccination— warrants further study as a correlate of within-host diversity independently of diversity associated with viral load or viral-load-related technical artefacts.

### Robustness of clinical associations to analytical thresholds

To evaluate how sensitive the associations discussed above were to the criteria used to identify minor variants, we generated three additional datasets with different levels of stringency across criteria. In Alternate Dataset 1, we used samples with 200x coverage or more over at least 98% of the genome. We required minor variants to have a minimum of 2% allele frequency (MAF) at sites with at least 200x coverage and be supported by a minimum of 100 reads. In Alternate Dataset 2, we used samples with 500x coverage or more over at least 98% of the genome, and required minor variants to have at least 3% MAF supported by a minimum of 20 reads. In Alternate Dataset 3, we used samples with 1000x coverage or more over at least 98% of the genome and minor variants with at least 1% MAF. Because the sample size was significantly reduced, we relaxed the Ct criterion from Ct<26 to Ct<35 for the regression analyses on vaccination status and healthcare worker surveillance, on the assumption that random errors in higher Ct samples would not differ between the groups. Despite the varying sample sizes and minor variant detection limits, all three datasets showed significantly higher minor variant richness in hospitalized patients (**SuppFig.6**). This factor was the most important in all three random forest models, it was in the top three highest coefficients in all three LASSO regression models in which all factors were included, and it had the highest coefficient in all three LASSO models in which temporally-biased factors were excluded. The other two factors of interest – vaccination status and healthcare worker status – differed significantly in the extent of the association depending on the dataset used. Minor variant richness was significantly lower in vaccinated patients and in healthcare workers in Alternate Dataset 1, but only the healthcare worker factor was significant in Alternate Dataset 2, and neither factor was significant in alternate dataset 3. The ROC AUC values for the random forest classification model were similar for all three datasets (0.58-0.60), while the fraction of variation explained by the LASSO regression model was slightly higher for alternate dataset 3 (28% for the model including all factors, 16% for the model excluding temporally biased factors). Aside from the hospitalization variable, the coefficients of many factors changed substantially between alternate datasets, even changing sign (for example, plasma treatment and monoclonal antibody treatment were associated with increased or decreased minor variant richness depending on the dataset). We concluded that the thresholds and criteria used to identify minor variants could significantly affect the strength of observed associations, but that the higher richness of minor variants in samples from patients requiring hospitalization was robust.

### Mutational patterns of highly prevalent minor variants

Having determined that minor variant richness was robustly associated with hospitalization, we set out to analyze the mutational patterns in the genome. Across all samples, minor variants were found mostly concentrated in the Orf8 and N genes, an observation consistent with previous characterization of these genes as hypermutable^18,19^ (**Fig.3a,b**); this enrichment pattern was similar in samples from hospitalized and non-hospitalized patients (**SuppFig. 7**). The most common within-host mutation observed was C>T, consistent with previous studies and with the hypothesis that nucleic acid editing by host enzymes contributes to the mutational spectrum^11^ (**Fig.3c**). Surprisingly, C>T mutations were also the most prevalent among non-reproducible minor variants, despite the fact that C>T mutations are not known to be common sequencing errors^20^ **(SuppFig. 8)**.

**Figure 3.**
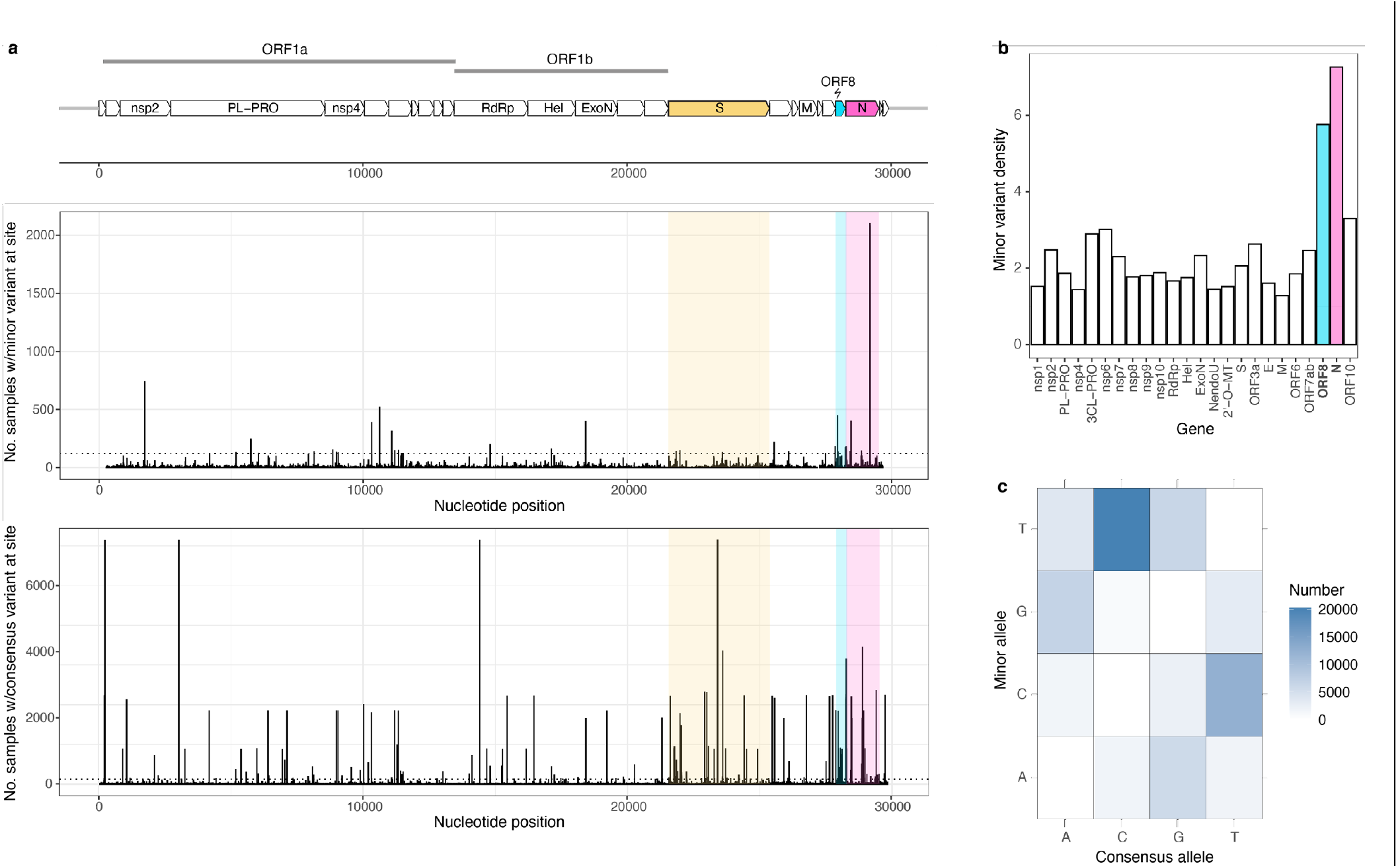
Genomic context of SARS-CoV-2 minor variants. **a**. Prevalence of minor variants (top) and consensus changes (bottom) at nucleotide positions across the genome. **b**. Density of minor variants in different SARS-CoV-2 genes (defined as total number of minor variants found in a gene across all samples divided by the length of the gene). **c**. Prevalence of different consensus to minor nucleotide substitutions.

We were particularly interested in sites containing minor variants in a high proportion of the samples. Mutational hotspots are of interest due to their potentially important role in convergent evolution. Therefore, a plausible purpose for monitoring minor variants in deep sequencing data is to identify sites with increased probabilities of mutation as a special focus for targeted mutational analysis. Doing so would require the ability to distinguish genuine mutational hotspots from recurrent artefacts.

We focused on 34 positions in the genome where minor variants were present in at least 2% of samples (**Fig.4a**). Minor variants at most of these positions were found at a range of frequencies from 1% to 50%, with several exceptions (**Fig.4b**). A majority of these sites included samples in which the minor variant was a reversion to the ancestral allele, suggesting repeated mutation at these sites. The gene containing the highest number (8) of highly recurrently mutated sites was N, but another 7 of these sites were found in the proteases PLpro or 3CLpro. These essential enzymes are involved in viral replication and immune modulation and thus high-profile targets for antiviral drug development^21,22^. This further justifies special attention to the mutational properties of these sites, since responsible development of antivirals ought to consider likely paths to the evolution of resistance, including loci with higher than average standing genetic variation within hosts. We cross-referenced the list of the 34 highly shared positions with highly shared sites from previous studies, with global patterns of consensus SNPs (queried from GISAID using cov-spectrum.org^23^), and with known phenotypically important convergent mutations^24^.

**Figure 4.**
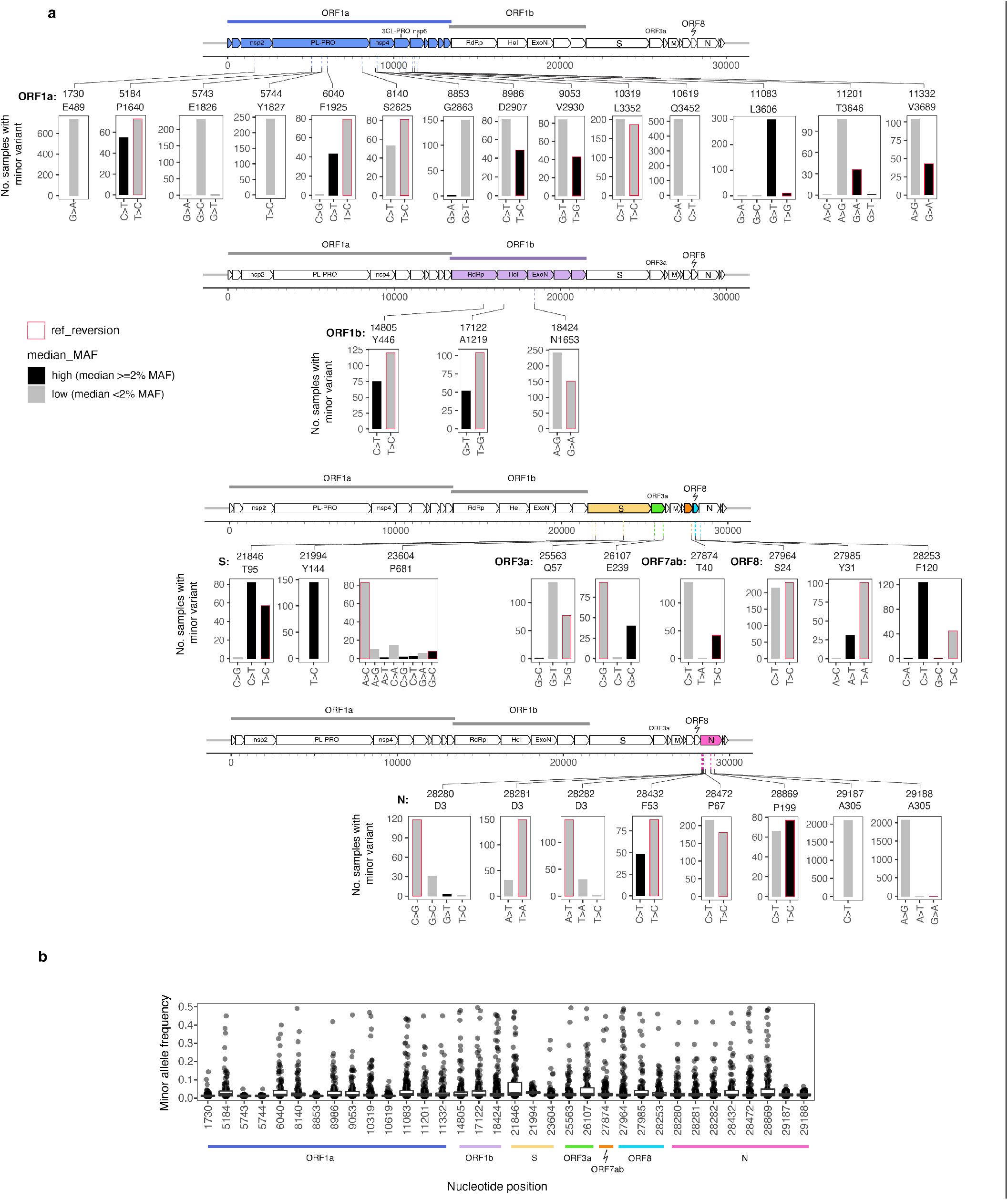
Nucleotide substitutions and minor allele frequencies at highly mutable sites. **a**. Minor variant changes at 34 nucleotide positions containing minor variants in at least 2% of samples. **b**. Minor allele frequencies of variants at these sites.

Several of the highly shared minor variant sites were highly polymorphic on the consensus level both within this dataset and in the US-wide GISAID data. Such sites pose challenges for interpretation because this pattern could be indicative of genuinely hypermutable sites but is also difficult to distinguish from cross-contamination because multiple consensus variants are often present in the same run. Indeed, in many sites that were polymorphic on the consensus level, minor variants only appeared in runs in which multiple consensus variants were present (**SuppFig.9**). One exception was nucleotide position 23604 (S: P681), a position proximal to the furin cleavage site that had all combinations of minor and consensus nucleotides present throughout the study period; and sites 28280-28282 (N: D3), which had two alleles at various frequencies present at each position in the codon throughout the study period. S: P681 has undergone different substitutions in multiple variants of concern; N: D3 contains a whole codon substitution in the Alpha lineage. These patterns are consistent with the explanation that these are highly mutable loci in which mutations have a high likelihood of becoming fixed in a lineage with a fitness advantage, like the variants of concern.

A somewhat puzzling pattern was observed for the highly prevalent minor variants at sites 1730 (orf1a: E489), 8853 (orf1a: G2863), 10619 (orf1a: Q3452), and 29187-29188 (N:A305) (**Fig.4a**). These sites were highly conserved at the consensus level but contained minor variants at low frequencies in a substantial fraction of specimens throughout the study period with a notable increase in the fraction of samples with minor variants in the later part of the study period (starting approximately with run 57/July 2021) (**SuppFig.9**). This period corresponds to when the vast majority of sequenced specimens were from the Delta lineages.

Some of the other sites had evidence suggesting technical or bioinformatic artefacts. For example, nucleotide position 11083 was identified as a highly recurrent minor variant in Lythgoe et al and Tonkin-Hill et all^10,11^, as well as a highly homoplasic site across the SARS-CoV-2 phylogeny^25^. It is immediately adjacent to a long T-homopolymer site, a well-known cause of sequencing error. Nucleotide position 21994 (S:Y144) is adjacent to a characteristic deletion in the Alpha (B.1.1.7) lineage, suggesting that mis-alignment of reads at the deletion site might contribute to the appearance of minor variants at that position. A particularly anomalous pair of minor variant sites were at positions 29187 and 29188, in the N gene, which were present in more than 30% of the samples studied but never at higher than 7% minor allele frequency. The amino acid mutation that these minor variants corresponded to, N: A305V, was extremely rare on the consensus level among sequences deposited to GISAID (**SuppFig.10)**, with only 72 sequences containing this mutation found in the United States, of which 59 were from Houston Methodist Hospital.

Finally, as another approach to examining whether convergent evolution could be linked to mutational patterns observed in minor variant data, we examined the frequency in the minor variant dataset of mutations repeatedly associated with increases in SARS-CoV-2 fitness. Obermeyer et al^24^ showed that single nucleotide mutations associated with increased transmissibility evolved independently multiple times in different lineages. Several mutations in the Spike protein, namely E484K, N501Y, K417N, and L18F, also independently evolved in several lineages of concern^26^. We examined the prevalence of these 22 nucleotide substitutions as minor variants in our data. Eight of these were not found as minor variants in any samples, whereas 14 (13 of them in the Spike protein) were found in at least one sample **(SuppFig.11)**. The most prevalent of these minor variant mutations was S:T95I, which was at nucleotide position 21846, one of the identified highly shared sites. This mutation, in the N-terminal domain of the Spike protein, has arisen independently in at least 30 consensus lineages and is associated with significant increases in viral fitness, but its phenotypic effect has not, to our knowledge, been experimentally characterized. Given the tendency of this mutation to frequently arise both within hosts and be successfully transmitted between hosts, further characterization of the effects of this change may be warranted to inform design of drugs and antibodies.

## Discussion

We explored the feasibility of characterizing within-host diversity by extracting minor variants data from clinical genomic surveillance samples of a large, densely sampled population to supplement consensus-level understanding of viral variants. The clearest finding of this study is that although within-host diversity is generally low, higher within-host diversity is associated with patients requiring hospitalization. Previous studies of minor variants in SARS-CoV-2 have consistently identified outlier samples with high numbers of within-host minor variants even after stringent quality control but were unable to examine the implications of such specimens due to their rarity. Exactly such rare events (i.e. patients with highly diverse viral populations) may be disproportionate drivers of viral recombination and transmission of standing genetic variation on which host-mediated natural selection can act. In this dataset, the absolute number of these outliers was high enough that we could test whether any of the host factors were associated with higher viral diversity.

Every novel consensus variant must at some point arise as a within-host variant, so it is crucial to understand what contexts may be likely to incubate viral population diversity. We found that, despite the noise introduced by variation in sequencing runs and sample Ct values, the signal was strong enough to observe a clear correlation between higher minor variant richness in more severely ill patients (those admitted to inpatient care or the ICU) and lower richness in vaccinated patients, in whom the number of replication cycles is assumed to be constrained. Previously, similar observations had been hinted at in smaller studies comparing cancer patients with healthcare workers, comparing mildly and severely ill patients, and examining minor variants in samples from patients of different ages ^27–29^. Studies in which patients were longitudinally sampled have shown fluctuating numbers of minor variants over time, with little directional trend^9,30^. Combined with our data, this suggests that within-host diversity is temporally dynamic, but in the aggregate is more likely to be high in more severely ill patients. This has implications for infection control strategies, for example bolstering the case that transmission control in healthcare settings and among symptomatically infected patients should be a critically high priority for preventing the emergence of new viral variants.

The direction of causality for the association between severe disease and high within-host diversity is unclear. It is plausible that more severe disease is the result of a more prolonged or quickly-replicating infection, during which more mutations can accumulate^31^; but, conversely, it is possible that more diverse infections drive more severe disease^32^. It is also possible that the immune responses during severe disease are distinctive in ways that affect selection on the viral population, or that there are mechanistic links between the comorbidities associated with risk of hospitalization and the dynamics of immune selection. For example, obesity is associated with more negative outcomes in influenza, and a study of influenza virus diversity in mice found that influenza A virus replicated faster and accumulated more diversity in obese than in lean mice, an effect that appeared to be mediated by the differential robustness of the interferon response ^33^. Future studies should conduct case-control comparisons of technically replicated longitudinal samples of patients with different disease time-courses to understand how virus population diversity changes over time in different types of hosts, under different immunological and clinical conditions. Such dynamics are well understood in viruses such as HIV, but may be more complex in SARS-CoV-2, which appears to elicit highly heterogeneous immune responses in different patients^34^.

In multiple previous studies of SARS-CoV-2 within-host diversity, mutational hotspots have hampered attempts to use minor variants to track transmission of closely related lineages^11,35^, since it is difficult to distinguish hypermutable sites, recurrent artefacts, and sites that are genuinely co-transmitted. Notably, in Tonkin-Hill et al, there was no correlation between the probability of transmission between two patients and the number of minor variants they shared; samples that were epidemiologically distant from each other often had more than 10 minor variants in common. This was true even after excluding minor variants that were generally highly prevalent in the dataset. Other authors have pointed out that automatically excluding minor variants that are present in many samples is not always warranted in epidemiological inference, for example when examining a superspreading event in which a large group of people were interacting and transmitting virus for a substantial length of time^35^. In general, it appears that convergent within-host *de novo* mutation is common enough to significantly complicate inferences of transmission of within-host diversity. For these reasons, it was warranted to play closer attention to the characteristics of sites containing minor variants in many samples. We found evidence both for highly recurrent artefacts and for phenotypically important recurrent mutations, the latter of which may be a high priority for targeted mutational studies.

Our observations suggest that identifying genuine mutational hotspots requires both understanding the genomic context (noting adjacent deletions, homopolymers or other structural features that might affect spurious minor variant calling) and also the wider sequencing context. For example, the increase in the prevalence of several mutations in the same later runs is difficult to explain. Although it is plausible that different lineages would have different within-host mutation rates, it is more difficult to imagine a mechanism by which certain lineages would have elevated rates of mutation only at specific sites. It is also difficult to rule out that some unknown technical change in sequencing conditions also contributed to changes in relative prevalence at these sites. Similarly, we suspect that the high prevalence of minor variants at sites 29187-29188 may be an artefact of the specific combination of methods used at this sequencing facility, because consensus variants at this position were very rare in consensus sequences from GISAID and primarily came from Houston Methodist, in samples with very different genetic backgrounds and collection dates. The existence of consensus mutations specific to particular sequencing labs was noted early in the pandemic^25^, so caution when detecting unusual mutations on the minor variant level is particularly warranted.

One of the purposes of this study was to examine the general feasibility of extracting minor variant data from samples not collected for this purpose. Despite the exceptional level of quality control involved in the generation of our sequences in the clinical context, we found that unavoidable technical artefacts, in particular batch effects and the clear effect of RNA input quantity on minor variant calling—even when limiting samples to those with high coverage across the genome—hampered the ability to draw definitive conclusions in the absence of technical replicates and required us to limit our analyses to high input samples. Our results demonstrate that caution is required when, for example, analyzing minor variants from sequence data repositories^36,37^. Rates of different types of error may meaningfully differ between batches of samples such as sequencing runs, between laboratory protocols and even due to factors such as whether individual tubes or plates were used in library preparation ^38,39^. If the idiosyncratic physical conditions under which library preparation occurs differentially affect error rates, then developing universally applicable error models may be extremely difficult. At the same time, the composition of batches may be non-random in biologically meaningful ways (e.g. samples from an outbreak in a particular location or population are likely to be sequenced in the same batch), making it difficult to disentangle biological and technical causes of patterns of minor variant prevalence. These results show that, without the development of more mature methods for correcting for numerous different sources of technical noise, deep sequence data cannot be used for routine monitoring of within-host viral diversity in the same way that consensus sequences are used for genomic surveillance.

Targeted studies of within-host diversity that take these technical issues into account can, however, lead to a greater understanding of the mutational biases of the virus and characteristics of the within-patient environment that affect viral diversification. In our exploratory study, the clearest emergent signal is that infections with high virus diversity are enriched among hospitalized patients. This has clear implications for prioritizing transmission control in healthcare settings and Further dissection of within-host viral dynamics is required to determine whether knowledge of a patient’s viral population diversity can better inform clinical care.

## Methods

### Patient population and ethics

The work was approved by the Houston Methodist Research Institute Institutional Review Board (IRB1010-0199). Specimens from patients were obtained primarily from symptomatic patients with a suspicion for COVID-19 disease from outpatient, emergency, labor and delivery, and other types of clinics. Specimens from healthcare workers were collected as part of a non-mandatory workplace surveillance program. Specimens were tested in the Molecular Diagnostics Laboratory at Houston Methodist Hospital using assays granted Emergency Use Authorization (EUA) from the FDA (https://www.fda.gov/medical-devices/emergency-situations-medical-devices/faqs-diagnostic-testing-sars-cov-2#offeringtests, last accessed June 7, 2021). Standardized specimen collection methods were used (https://vimeo.com/396996468/2228335d56,). Multiple molecular testing platforms were used, including the COVID-19 test or RP2.1 test with BioFire Film Array instruments, the Xpert Xpress SARS-CoV-2 test using Cepheid GeneXpert Infinity or Cepheid GeneXpert Xpress IV instruments, the Cobas SARS-CoV-2 & Influenza A/B Assay using the Roche Liat system, the SARS-CoV-2 Assay using the Hologic Panther instrument, the Aptima SARS-CoV-2 Assay using the Hologic Panther Fusion system, the Cobas SARS-CoV-2 test using the Roche 6800 system, and the SARS-CoV-2 assay using Abbott Alinity instruments.

### Library preparation and sequencing

Libraries for whole SARS-CoV-2 genome sequencing were prepared according to version 3 (https://community.artic.network/t/sars-cov-2-version-4-scheme-release/312, last accessed August 19, 2021) of the ARTIC nCoV-2019 sequencing protocol. We used a semi-automated workflow described previously^6,7^ that employed BioMek i7 liquid handling workstations (Beckman Coulter Life Sciences) and MANTIS automated liquid handlers (FORMULATRIX). Short sequence reads were generated with a NovaSeq 6000 instrument (Illumina).

### Sample selection

The initial dataset was comprised of 39,880 samples from 70 Novaseq runs. These runs also often contained samples from other institutions or collection time periods, which we took into account when assessing the possibility of within-run cross-contamination but did not otherwise analyze. Median sequence depth of samples was broadly but not perfectly correlated with sample input quantity. We noted that very low input samples, i.e. with Ct values >=40, generally had very low coverage, but there was a small subset with high coverage comparable to high input samples (**SuppFig.12**). We excluded from the analysis any runs containing at least three Ct>=40 samples in which the coverage breadth and depth were not statistically different from the Ct<40 samples in the run (t-test>0.01 for median coverage or for fraction of genome with at least 1000x coverage). We further limited all our analyses to samples with at least 100x coverage over at least 98% of the genome excluding the 5’ and 3’ UTRs. We also excluded samples where the consensus sequence was flagged as poor quality under the default quality control criteria of Nextclade^15^ (QC score >100) or that did not have a lineage assigned by Pango^16^. In cases where multiple samples were collected longitudinally from the same patient, we chose the earliest sample.

### Consensus sequence assembly and minor variant calling

Adapter sequences were trimmed from reads using trimmomatic v0.39^40^ with the following options: ILLUMINACLIP:${params.adapters}:2:30:10:8:true LEADING:20 TRAILING:20 SLIDINGWINDOW:4:20 MINLEN:20. Reads were aligned to the Wuhan/Hu-1 SARS-CoV2 genome (RefSeq: NC_045512.2) using minimap2 v2.17 with the preset genomic short-read mapping option^41^. ARTIC v3 primer sequences^42^ were removed using iVar v.1.3.1 with a minimum quality threshold of 0 and including all reads with no primer sequences found^43^. Consensus sequences and minor variants were called using an in-house variant calling pipeline, timo, available at https://github.com/GhedinLab/timo.

### Analysis

Calculations, visualizations and statistical analyses were carried out in R v4.0.3 (R Foundation for Statistical Computing). Packages used for analysis included tidyverse 1.3.1 ^44^, glmnet 4.1.2 ^45^, nlme 3.1.149 ^46^, randomForest 4.6.14 ^47^, pROC 1.17.0.1. Consensus sequence quality control was carried out in Nextclade 1.4.1 ^15^.

### Additional Data Files

Inclusion_table.csv includes IDs of samples with information about which samples were included in which analyses, and will include SRA accession numbers for each sample when data deposition is complete. Files used in minor variant analysis are available in Github repository https://github.com/GhedinSGS/HMH-SARS-CoV2-minorvariants.

### Sequence and code availability

Raw sequence data are available under Bioproject PRJNA767338. Pipeline used for minor variant calling is available at https://github.com/GhedinLab/timo, and data files and code used for analyses are available at https://github.com/GhedinSGS/HMH-SARS-CoV2-minorvariants.

## Acknowledgements

We thank Drs. Marc Boom and Dirk Sostman for their ongoing support. We thank Matthew Ojeda Saavedra, Kristina Reppond, Madison N Shyer, Jessica Cambric, Ryan Gadd, Rashi M Thakur, Akanksha Batajoo, Regan Mangham, Sindy Pena and Trina Trinh for technical support. The research was supported by the Houston Methodist Academic Institute Infectious Diseases Fund and many generous Houston philanthropists. This work utilized the computational resources of the NIH High Performance Computing (HPC) Biowulf cluster (http://hpc.nih.gov).

## Funding Statement

This project was supported in part by the Houston Methodist Academic Institute Infectious Diseases Fund, and federal funds from the National Institute of Allergy and Infectious Diseases, National Institutes of Health, Department of Health and Human Services, under Contract No. 75N93019C00076 (J.J.D.). This work was also supported in part by the Division of Intramural Research (DIR) of the NIAID/NIH.

## Supplemental Figures

**Supplementary Figure 1. Reproducibility of minor variants in 54 samples with technical replicates. a,b**. Reproducibility of minor variant detection (black vs. red) and minor allele frequency in samples with different input quantities, by category and by individual samples. “Unknown” Ct values represent samples diagnosed on the Hologic Aptima instrument, which gives results in relative light units (values >100 in individual sample panels) or on the Biofire Diagnostics instrument, which does not quantify viral load (“NA” in individual sample panels). **c**. Minor variant reproducibility in samples with different ranges of median sequencing depth. **d**. Sequencing depth at site and minor allele frequency of minor variants that were subsequently detected in the second technical replicate (“yes”) vs. not detected (“no”).

**Supplementary Figure 2. Collection dates and consensus virus lineage of final sample set**.

**Supplementary Figure 3. Run-level effects on minor variant richness in filtered sample set. a**. Minor variant richness in high coverage and Ct<26 samples in each sequencing run. **b**. Relationship between sample’s median sequencing coverage and number of minor variants. **c**. Median number of minor variants for samples in each run represented as a function of run-level median of median coverage.

**Supplementary Figure 4. Categories of minor variant diversity among hospitalized and non-hospitalized patients**.

**Supplementary Figure 5. LASSO regression coefficients for association of patient and sample factors of interest with minor variant richness. a**. Results of model in which all factors of interest are included. **b**. Results of model in which factors with a strong temporal dimension were excluded (collection month, consensus lineage, vaccination status).

**Supplementary Figure 6. Associations between patient status and SARS-CoV-2 diversity in three datasets using different thresholds for sample coverage and minor variant detection**.

**Supplementary Figure 7. Density of minor variants in different SARS-CoV-2 genes in samples from hospitalized and non-hospitalized patients**. Minor variant density is defined as total number of minor variants found in a gene across all samples divided by the length of the gene.

**Supplementary Figure 8**. Prevalence of different consensus > minor nucleotide substitutions at minor variant sites that were detected vs. not detected in a second sequencing replicate.

**Supplementary Figure 9. Prevalence of minor alleles and consensus alleles at the 34 most frequent minor variant sites, by run**. For consensus allele prevalence, all samples from the run are included, regardless of whether they were analyzed in the minor variant study, on the assumption that they may contribute to cross-contamination in other samples.

**Supplementary Figure 10. Minor variant prevalence in the Houston Methodist dataset vs. prevalence of consensus mutations at these sites in US-wide SARS-CoV-2 sequences from GISAID**. GISAID sequences were queried on June 13, 2022 covering the entire length of the pandemic in the U.S. to identify the number of sequences that had any nucleotide substitution (A,C,T,G) at the 34 nucleotide positions that most frequently had minor variants in the Houston dataset.

**Supplementary Figure 11. Prevalence across time of minor variants corresponding to recurrent amino acid changes associated with increased SARS-CoV-2 fitness**, as identified in Obermeyer et al^24^.

**Supplementary Figure 12. Sequencing depth and Ct values of Houston Methodist SARS-CoV-2 samples prior to filtering**.

## Notes

### Competing Interest Statement

The authors have declared no competing interest.

### Author Declarations

The work was approved by the Houston Methodist Research Institute Institutional Review Board (IRB1010-0199)

## References

1. du Plessis, L. et al. Establishment and lineage dynamics of the SARS-CoV-2 epidemic in the UK. Science eabf2946 (2021) doi:10.1126/science.abf2946.

2. Grubaugh, N. D. et al. An amplicon-based sequencing framework for accurately measuring intrahost virus diversity using PrimalSeq and iVar. Genome Biol. 20, 8 (2019).

3. Worby, C. J., Lipsitch, M. & Hanage, W. P. Shared Genomic Variants: Identification of Transmission Routes Using Pathogen Deep-Sequence Data. Am. J. Epidemiol. 186, 1209–1216 (2017).

4. The CITIID-NIHR BioResource COVID-19 Collaboration et al. SARS-CoV-2 evolution during treatment of chronic infection. Nature 592, 277–282 (2021).

5. Roder, Ae. et al. Diversity and selection of SARS-CoV-2 minority variants in the early New York City outbreak. http://biorxiv.org/lookup/doi/10.1101/2021.05.05.442873 (2021) doi:10.1101/2021.05.05.442873.

6. Long, S. W. et al. Sequence Analysis of 20,453 Severe Acute Respiratory Syndrome Coronavirus 2 Genomes from the Houston Metropolitan Area Identifies the Emergence and Widespread Distribution of Multiple Isolates of All Major Variants of Concern. Am. J. Pathol. 191, 983–992 (2021).

7. Olsen, R. J. et al. Trajectory of Growth of Severe Acute Respiratory Syndrome Coronavirus 2 (SARS-CoV-2) Variants in Houston, Texas, January through May 2021, Based on 12,476 Genome Sequences. Am. J. Pathol. 191, 1754–1773 (2021).

8. Christensen, P. A. et al. Signals of Significantly Increased Vaccine Breakthrough, Decreased Hospitalization Rates, and Less Severe Disease in Patients with Coronavirus Disease 2019 Caused by the Omicron Variant of Severe Acute Respiratory Syndrome Coronavirus 2 in Houston, Texas. Am. J. Pathol. S000294402200044X (2022) doi:10.1016/j.ajpath.2022.01.007.

9. Valesano, A. L. et al. Temporal dynamics of SARS-CoV-2 mutation accumulation within and across infected hosts. PLOS Pathog. 17, e1009499 (2021).

10. Lythgoe, K. A. et al. SARS-CoV-2 within-host diversity and transmission. Science eabg0821 (2021) doi:10.1126/science.abg0821.

11. Tonkin-Hill, G. et al. Patterns of within-host genetic diversity in SARS-CoV-2. eLife 10, e66857 (2021).

12. Braun, K. M. et al. Acute SARS-CoV-2 infections harbor limited within-host diversity and transmit via tight transmission bottlenecks. PLOS Pathog. 17, e1009849 (2021).

13. Storz, J. F. et al. The role of mutation bias in adaptive molecular evolution: insights from convergent changes in protein function. 1.

14. Stoltzfus, A. & Yampolsky, L. Y. Climbing Mount Probable: Mutation as a Cause of Nonrandomness in Evolution. J. Hered. 100, 637–647 (2009).

15. Hadfield, J. et al. Nextstrain: real-time tracking of pathogen evolution. Bioinformatics 34, 4121–4123 (2018).

16. O’Toole, Á. et al. Assignment of Epidemiological Lineages in an Emerging Pandemic Using the Pangolin Tool. Virus Evol. veab064 (2021) doi:10.1093/ve/veab064.

17. Ortiz, A. T. et al. Within-host diversity improves phylogenetic and transmission reconstruction of SARS-CoV-2 outbreaks. http://biorxiv.org/lookup/doi/10.1101/2022.06.07.495142 (2022) doi:10.1101/2022.06.07.495142.

18. Zinzula, L. Lost in deletion: The enigmatic ORF8 protein of SARS-CoV-2. Biochem. Biophys. Res. Commun. 538, 116–124 (2021).

19. Lin, M. J. et al. Host–pathogen dynamics in longitudinal clinical specimens from patients with COVID-19. Sci. Rep. 12, 5856 (2022).

20. Stoler, N. & Nekrutenko, A. Sequencing error profiles of Illumina sequencing instruments. NAR Genomics Bioinforma. 3, lqab019 (2021).

21. Rut, W. et al. Activity profiling and crystal structures of inhibitor-bound SARS-CoV-2 papain-like protease: A framework for anti-COVID-19 drug design. Sci. Adv. 6, eabd4596 (2020).

22. Shin, D. et al. Papain-like protease regulates SARS-CoV-2 viral spread and innate immunity. Nature 587, 657–662 (2020).

23. Chen, C. et al. CoV-Spectrum: analysis of globally shared SARS-CoV-2 data to identify and characterize new variants. Bioinformatics 38, 1735–1737 (2022).

24. Obermeyer, F. et al. Analysis of 6.4 million SARS-CoV-2 genomes identifies mutations associated with fitness. Science abm1208 (2022) doi:10.1126/science.abm1208.

25. de Maio, N. et al. Issues with SARS-CoV-2 sequencing data.

26. Martin, D. P. et al. The emergence and ongoing convergent evolution of the SARS-CoV-2 N501Y lineages. Cell S0092867421010503 (2021) doi:10.1016/j.cell.2021.09.003.

27. Siqueira, J. D. et al. SARS-CoV-2 genomic and quasispecies analyses in cancer patients reveal relaxed intrahost virus evolution. http://biorxiv.org/lookup/doi/10.1101/2020.08.26.267831 (2020) doi:10.1101/2020.08.26.267831.

28. Al Khatib, H. A. et al. Within-Host Diversity of SARS-CoV-2 in COVID-19 Patients With Variable Disease Severities. Front. Cell. Infect. Microbiol. 10, 575613 (2020).

29. Kuipers, J. et al. Within-patient genetic diversity of SARS-CoV-2. http://biorxiv.org/lookup/doi/10.1101/2020.10.12.335919 (2020) doi:10.1101/2020.10.12.335919.

30. Simons, L. M. et al. Assessment of Virological Contributions to COVID-19 Outcomes in a Longitudinal Cohort of Hospitalized Adults. Open Forum Infect. Dis. 9, ofac027 (2022).

31. Li, J. et al. Two-step fitness selection for intra-host variations in SARS-CoV-2. Cell Rep. 110205 (2021) doi:10.1016/j.celrep.2021.110205.

32. Töpfer, A. et al. Sequencing approach to analyze the role of quasispecies for classical swine fever. Virology 438, 14–19 (2013).

33. Honce, R. et al. Obesity-Related Microenvironment Promotes Emergence of Virulent Influenza Virus Strains. mBio 11, (2020).

34. Mathew, D. et al. Deep immune profiling of COVID-19 patients reveals distinct immunotypes with therapeutic implications. Science 369, eabc8511 (2020).

35. Nicholson, M. D. et al. Response to comment on “Genomic epidemiology of superspreading events in Austria reveals mutational dynamics and transmission properties of SARS-CoV-2”. Sci. Transl. Med. 13, eabj3222 (2021).

36. Pathak, A. K. et al. Spatio-temporal dynamics of intra-host variability in SARS-CoV-2 genomes. 11.

37. Armero, A., Berthet, N. & Avarre, J.-C. Intra-Host Diversity of SARS-Cov-2 Should Not Be Neglected: Case of the State of Victoria, Australia. Viruses 13, 133 (2021).

38. Walker, A. W. A Lot on Your Plate? Well-to-Well Contamination as an Additional Confounder in Microbiome Sequence Analyses. mSystems 4, (2019).

39. Lam, C. et al. Sars-CoV-2 Genome Sequencing Methods Differ In Their Ability To Detect Variants From Low Viral Load Samples. J. Clin. Microbiol. (2021) doi:10.1128/JCM.01046-21.

40. Bolger, A. M., Lohse, M. & Usadel, B. Trimmomatic: a flexible trimmer for Illumina sequence data. Bioinformatics 30, 2114–2120 (2014).

41. Li, H. Minimap2: pairwise alignment for nucleotide sequences. Bioinformatics 34, 3094–3100 (2018).

42. Tyson, J. R. et al. Improvements to the ARTIC multiplex PCR method for SARS-CoV-2 genome sequencing using nanopore. http://biorxiv.org/lookup/doi/10.1101/2020.09.04.283077 (2020) doi:10.1101/2020.09.04.283077.

43. Castellano, S. et al. iVar, an Interpretation-Oriented Tool to Manage the Update and Revision of Variant Annotation and Classification. Genes 12, 384 (2021).

44. Wickham, H. et al. Welcome to the Tidyverse. J. Open Source Softw. 4, 1686 (2019).

45. Friedman, J., Hastie, T. & Tibshirani, R. Regularization Paths for Generalized Linear Models via Coordinate Descent. J. Stat. Softw. 33, (2010).

46. nlme: Linear and Nonlinear Mixed Effects Models.

47. Liaw, A. & Wiener, M. Classification and Regression by randomForest. 2, 5 (2002).

