## Supplemental Figures for "Within-host genetic diversity of SARS-CoV-2 in the context of large-scale hospital-associated genomic surveillance"

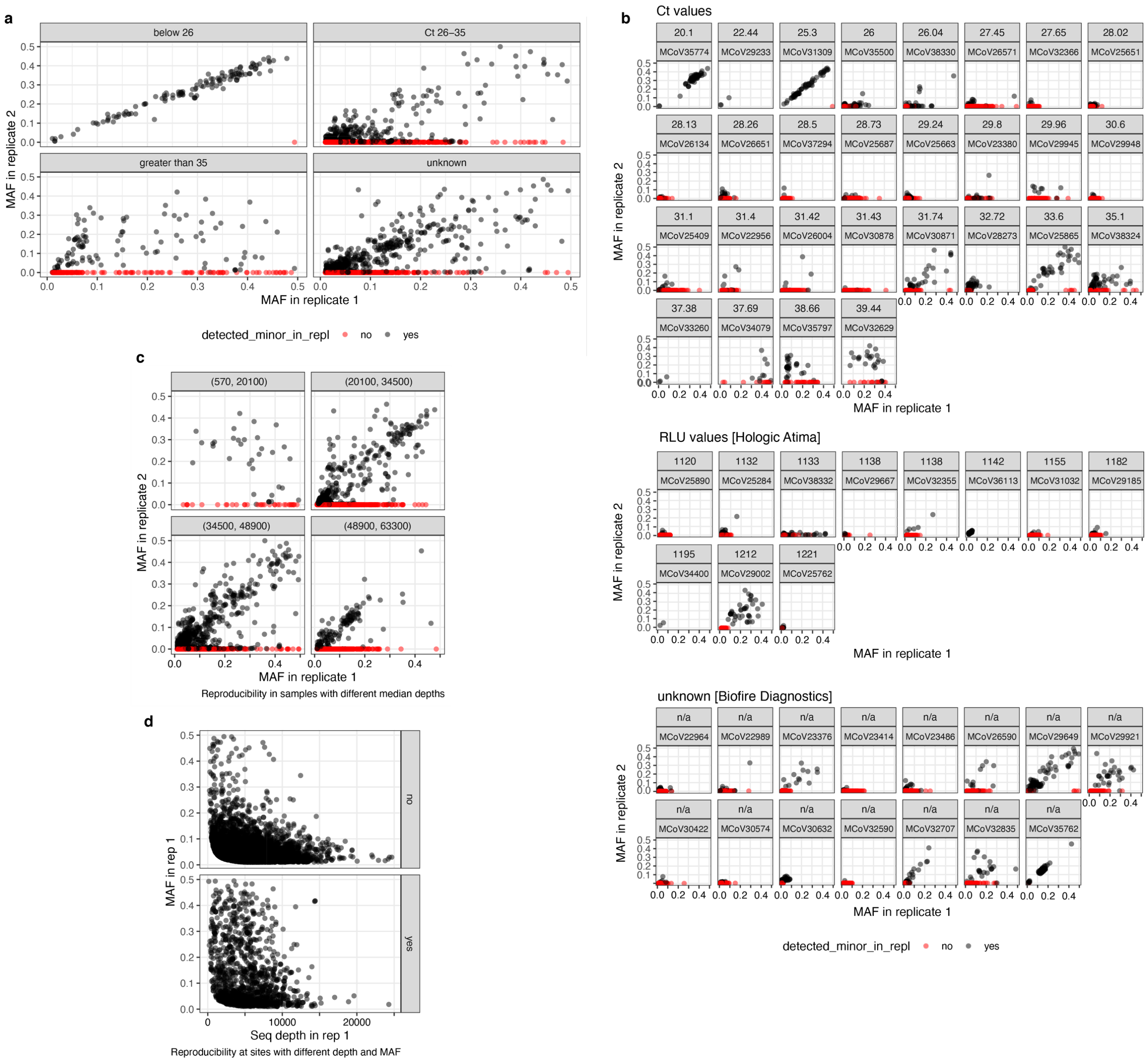


**Supplementary Figure 1. Reproducibility of minor variants in 54 samples with technical replicates. a,b**. Reproducibility of minor variant detection (black vs. red) and minor allele frequency in samples with different input quantities, by category and by individual samples. "Unknown" Ct values represent samples diagnosed on the Hologic Aptima instrument, which gives results in relative light units (values >100 in individual sample panels) or on the Biofire Diagnostics instrument, which does not quantify viral load ("NA" in individual sample panels). **c**. Minor variant reproducibility in samples with different ranges of median sequencing depth. **d**. Sequencing depth at site and minor allele frequency of minor variants that were subsequently detected in the second technical replicate ("yes") vs. not detected ("no").

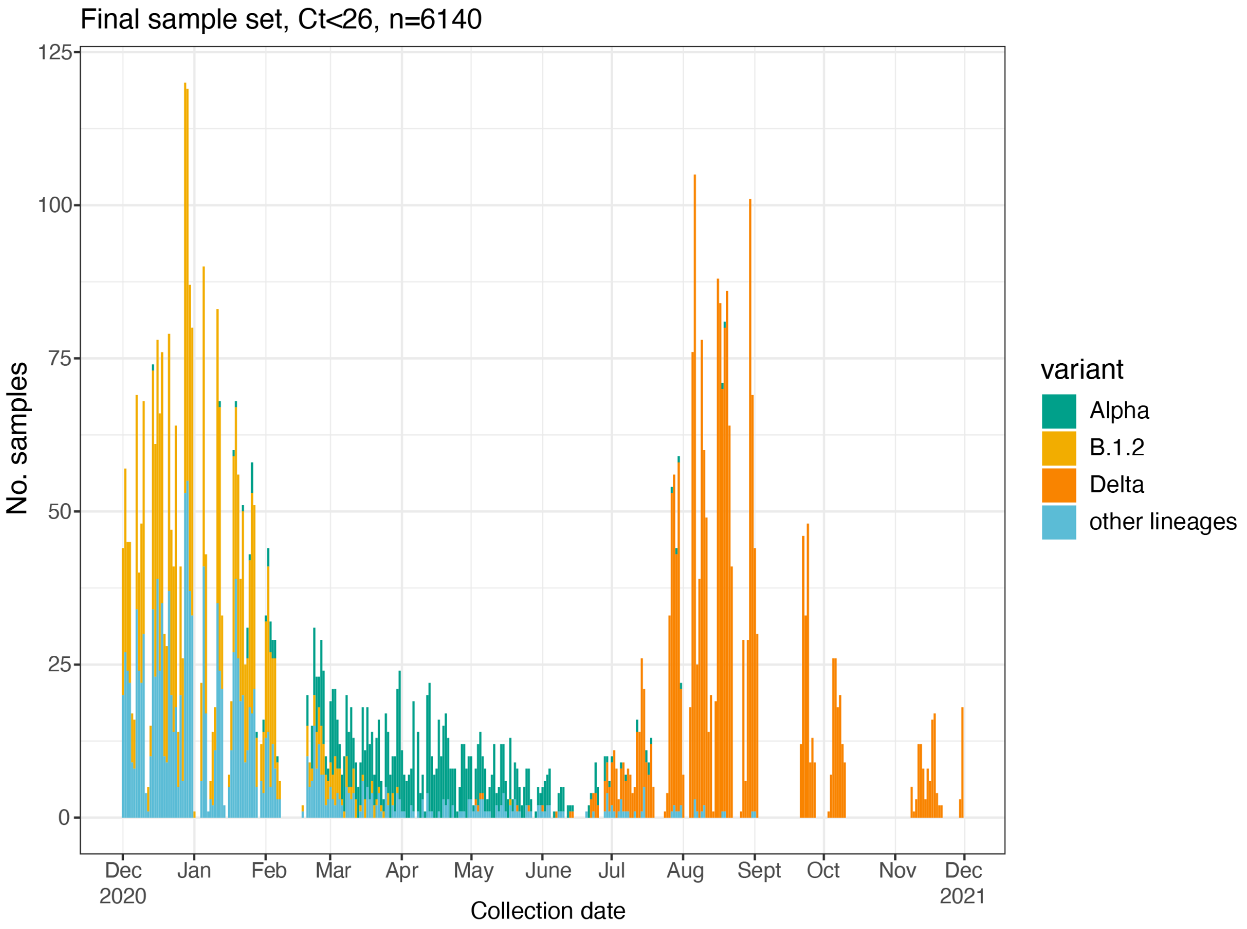


**Supplementary Figure 2. Collection dates and consensus virus lineage of final sample set.**

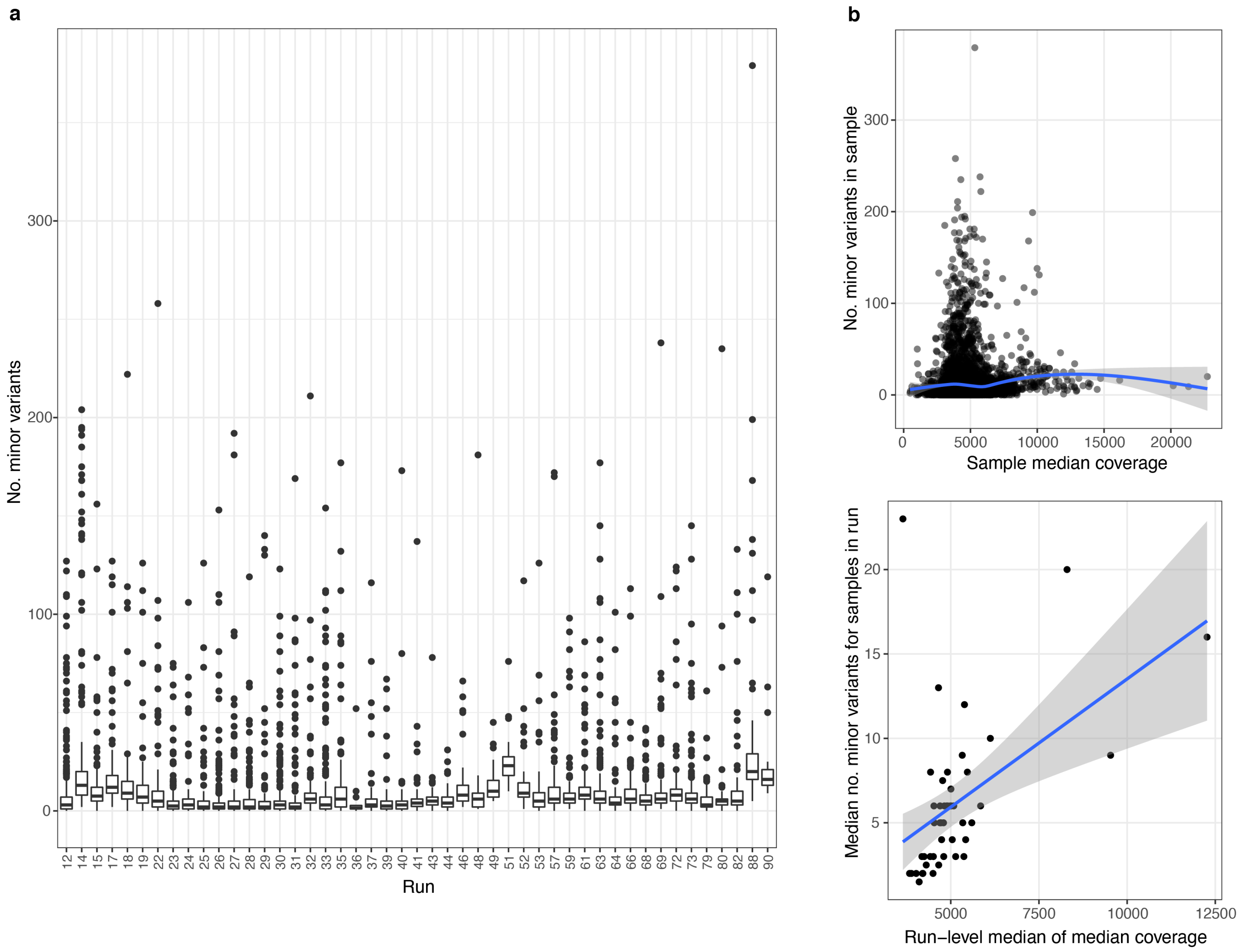


**Supplementary Figure 3. Run-level effects on minor variant richness in filtered sample set. a**. Minor variant richness in high coverage and Ct<26 samples in each sequencing run. **b.** Relationship between sample's median sequencing coverage and number of minor variants. **c.** Median number of minor variants for samples in each run represented as a function of run–level median of median coverage.

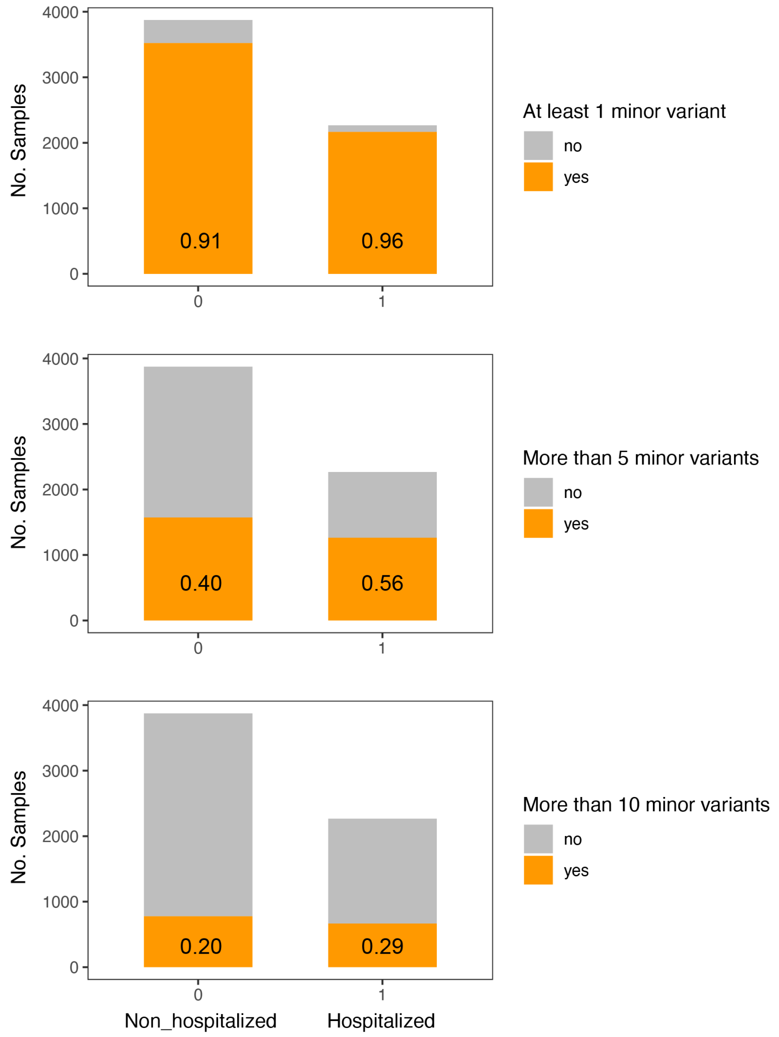


**Supplementary Figure 4. Categories of minor variant diversity among hospitalized and non-hospitalized patients.**

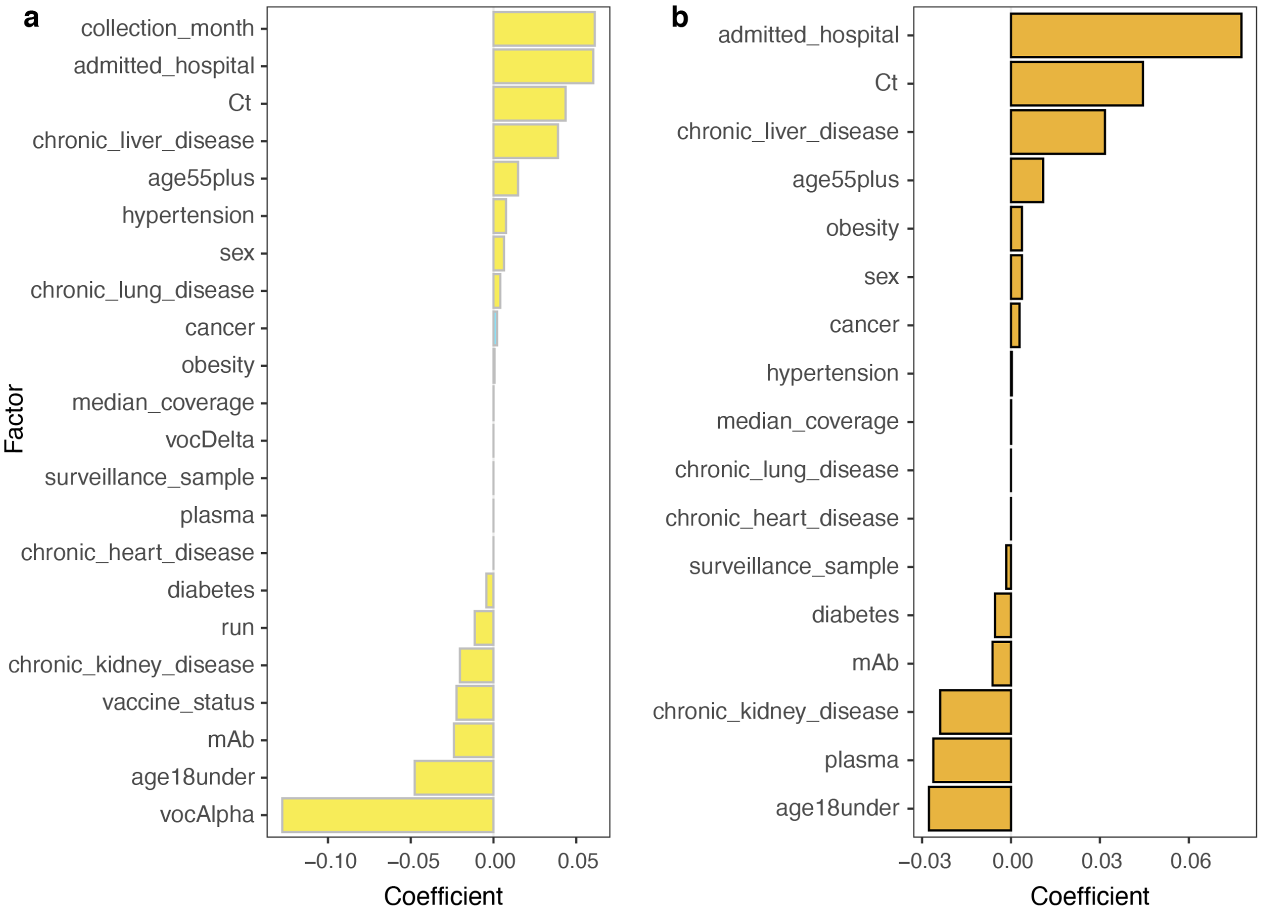


**Supplementary Figure 5. LASSO regression coefficients for association of patient and sample factors of interest with minor variant richness. a.** Results of model in which all factors of interest are included. **b.** Results of model in which factors with a strong temporal dimension were excluded (collection month, consensus lineage, vaccination status).

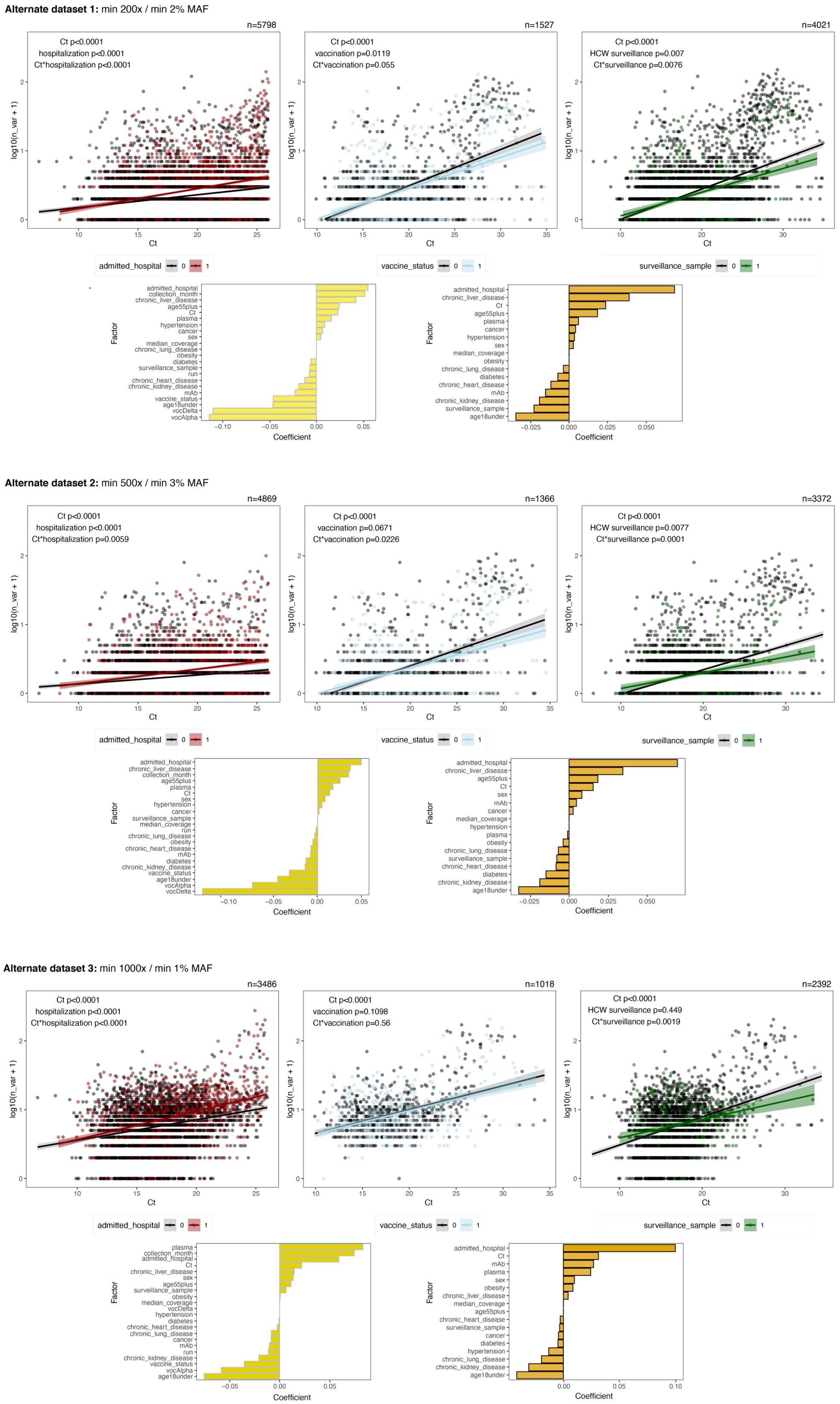


**Supplementary Figure 6. Associations between patient status and SARS-CoV-2 diversity in three datasets using different thresholds for sample coverage and minor variant detection.**

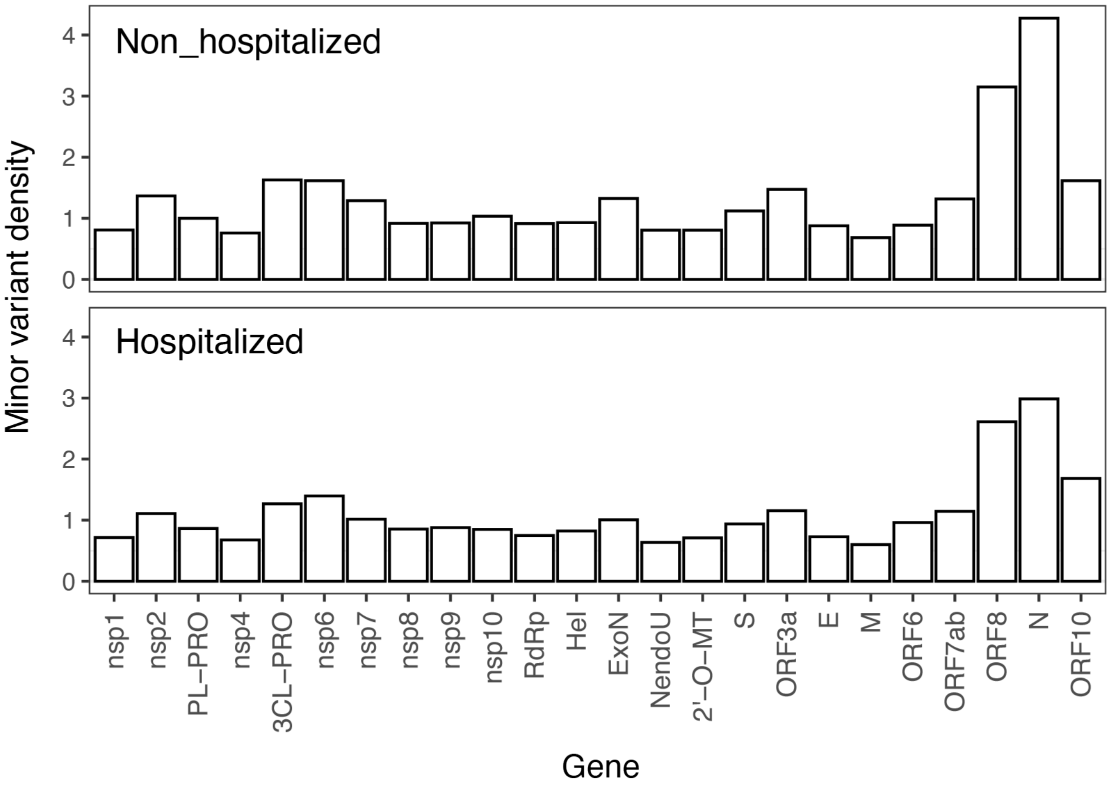


**Supplementary Figure 7. Density of minor variants in different SARS-CoV-2 genes in samples from hospitalized and non-hospitalized patients.** Minor variant density is defined as total number of minor variants found in a gene across all samples divided by the length of the gene.

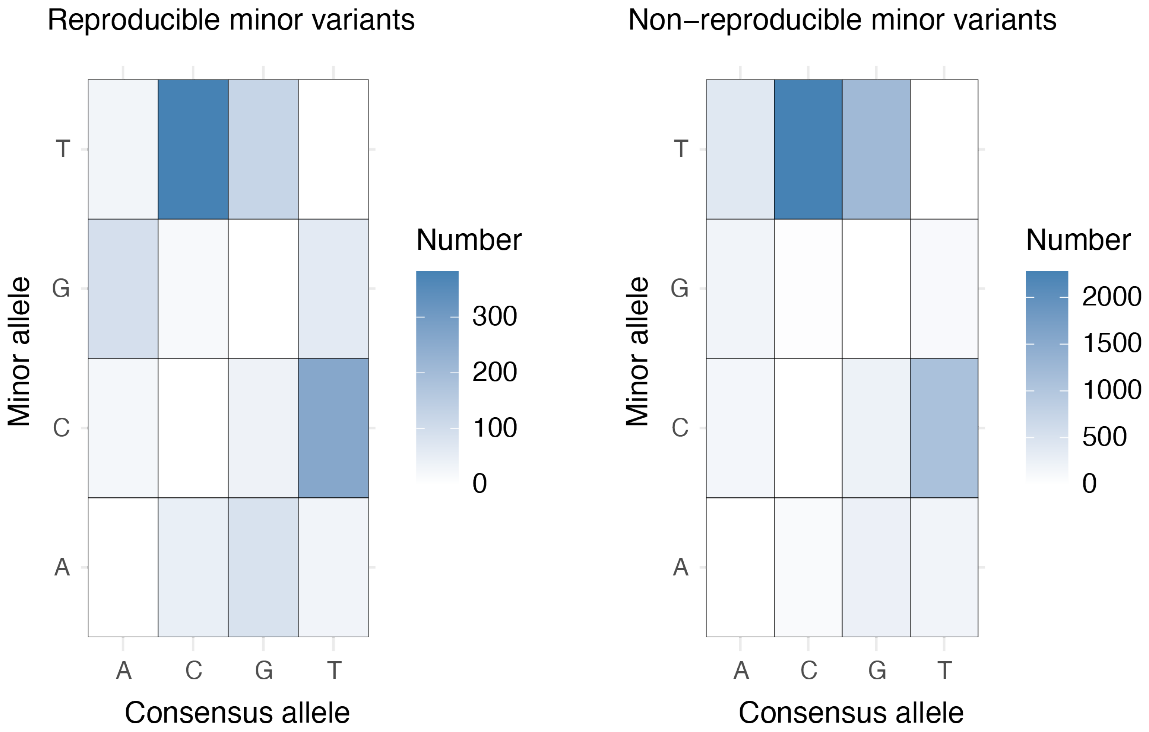


**Supplementary Figure 8.** Prevalence of different consensus > minor nucleotide substitutions at minor variant sites that were detected vs. not detected in a second sequencing replicate.

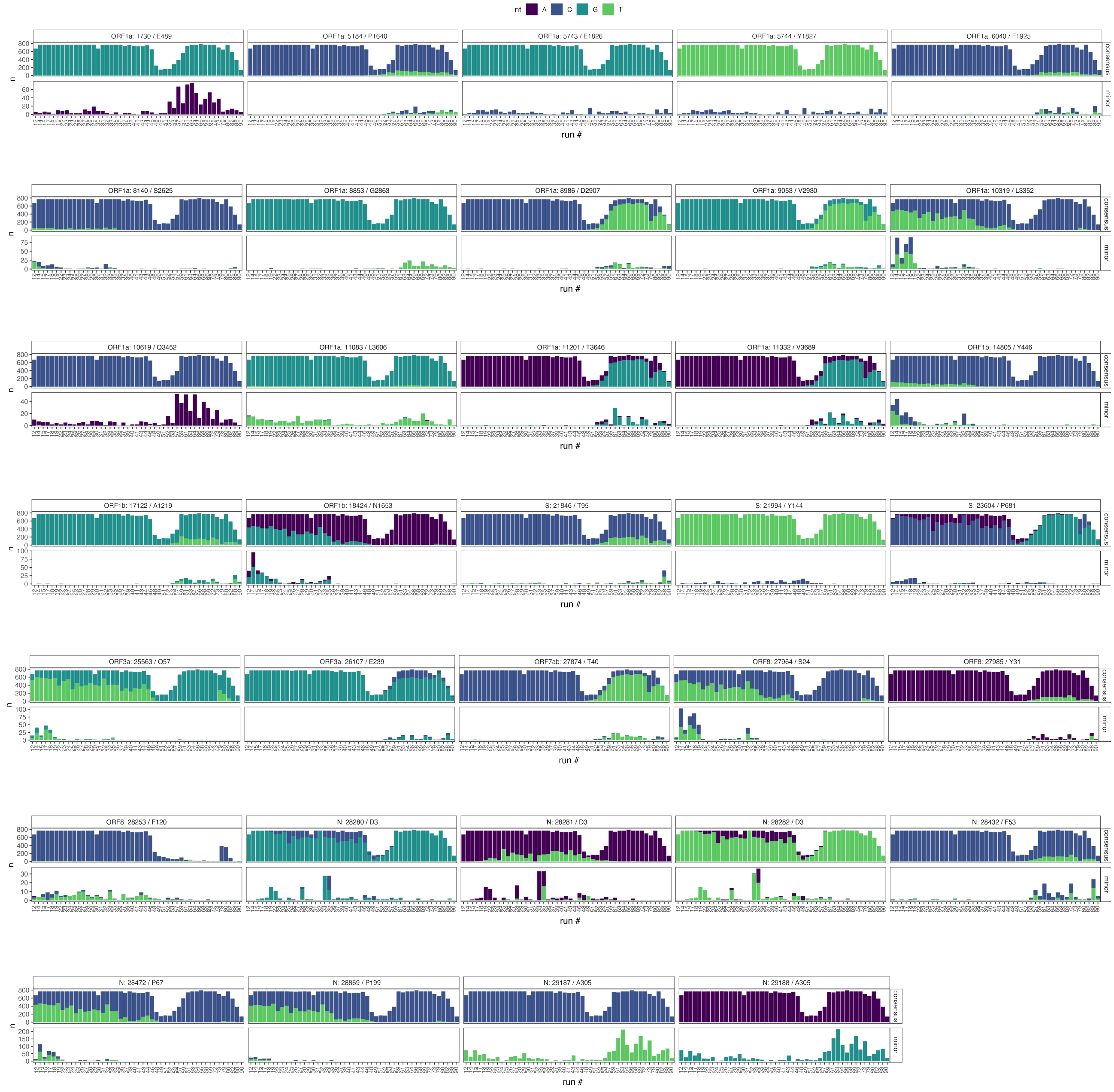


**Supplementary Figure 9. Prevalence of minor alleles and consensus alleles at the 34 most frequent minor variant sites, by run**. For consensus allele prevalence, all samples from the run are included, regardless of whether they were analyzed in the minor variant study, on the assumption that they may contribute to cross-contamination in other samples.

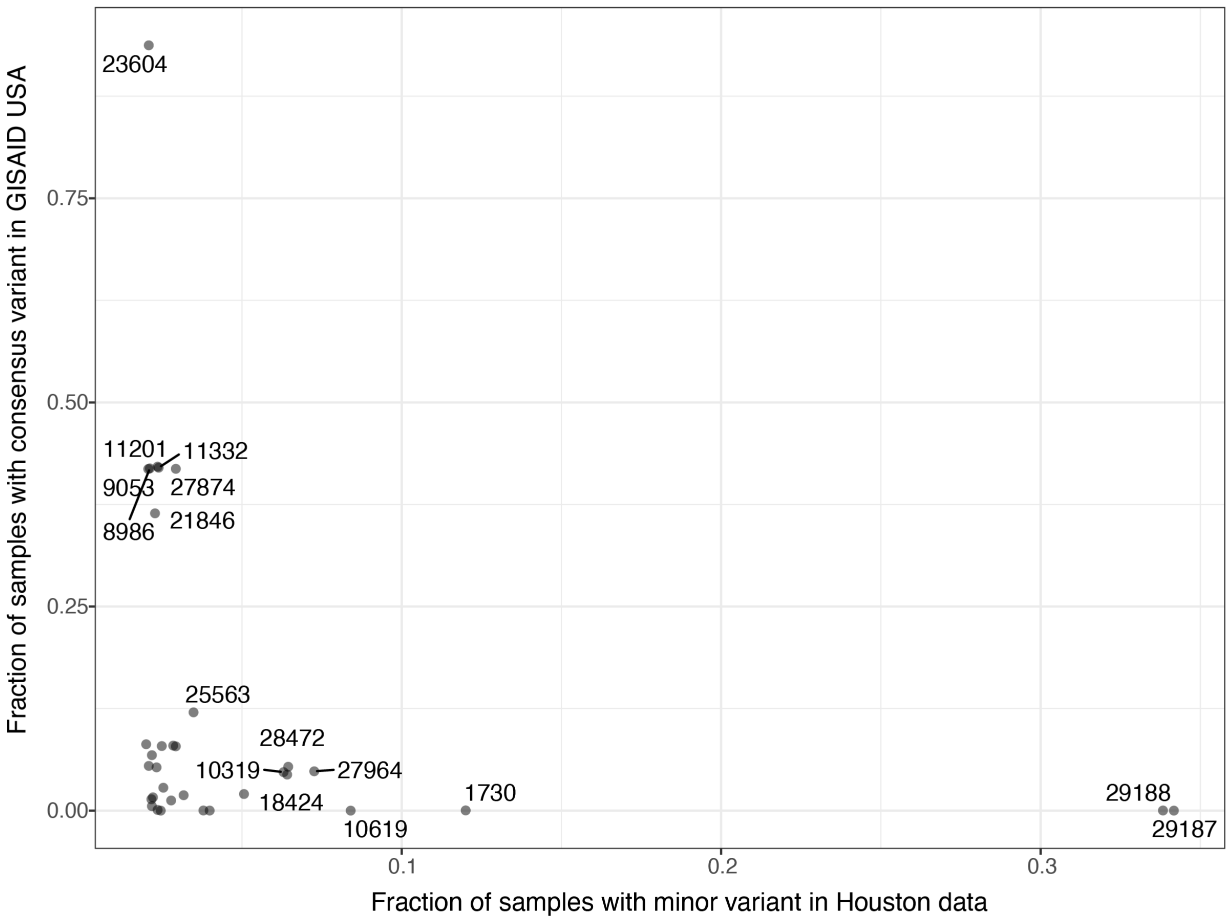


**Supplementary Figure 10. Minor variant prevalence in the Houston Methodist dataset vs. prevalence of consensus mutations at these sites in US-wide SARS-CoV-2 sequences from GISAID.** GISAID sequences were queried on June 13, 2022 covering the entire length of the pandemic in the U.S. to identify the number of sequences that had any nucleotide substitution (A,C,T,G) at the 34 nucleotide positions that most frequently had minor variants in the Houston dataset.

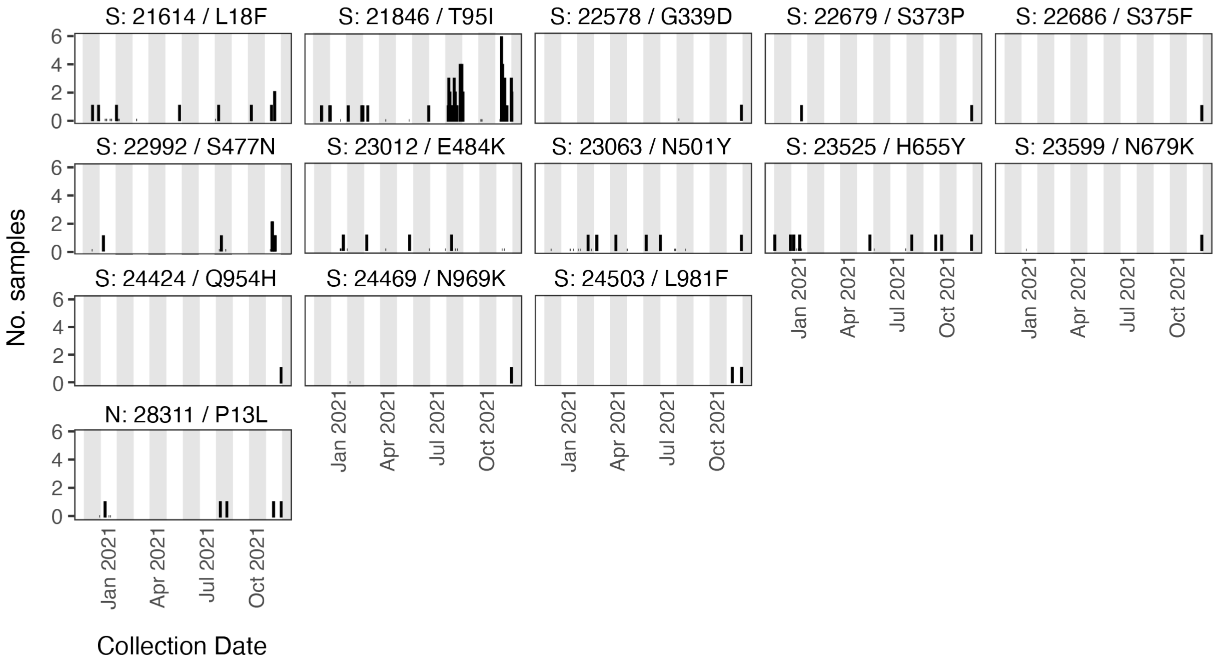


**Supplementary Figure 11. Prevalence across time of minor variants corresponding to recurrent amino acid changes associated with increased SARS-CoV-2 fitness**, as identified in Obermeyer et al[24].

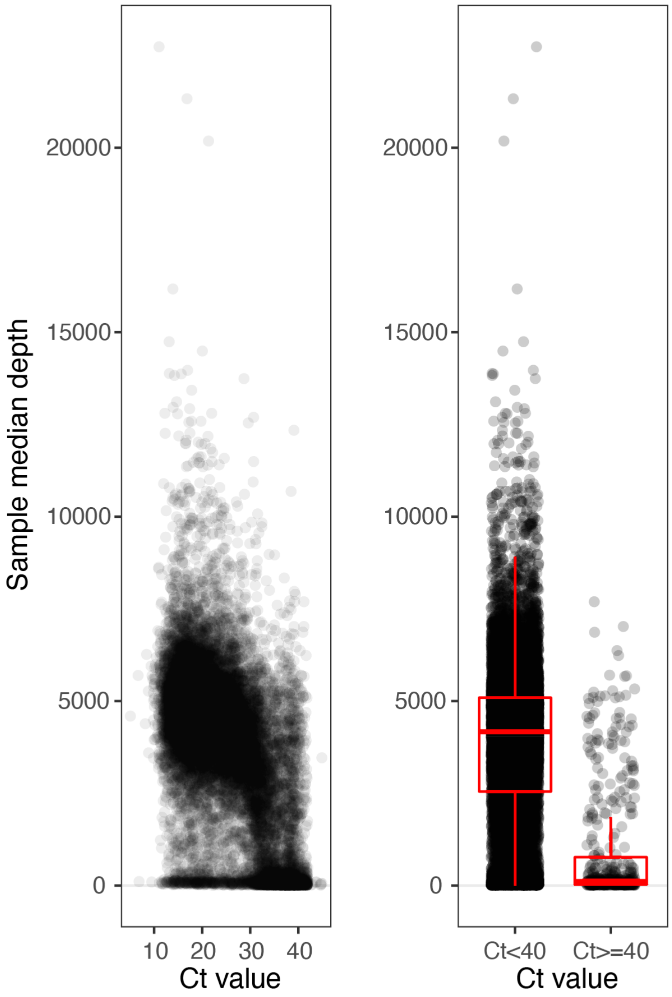


**Supplementary Figure 12. Sequencing depth and Ct values of Houston Methodist SARS-CoV-2 samples prior to filtering.**
